# Extracellular alpha satellite DNA in human plasma as a novel molecular biomarker for early diagnosis of bladder cancer

**DOI:** 10.1101/2025.02.14.25322062

**Authors:** D. Đermić, I. Feliciello

## Abstract

Although bladder cancer (BC) is a common urological disease, there are deficiencies in current methods used for its detection, hence the research on the identification of non-invasive biomarkers is given a high priority. Alpha satellite DNA is a major human satellite DNA (hASAT), which is important for genome stability and whose aberrant overexpression is a hallmark of many human cancers. Here we identified and quantified its release into the circulation of BC patients, demonstrating its increased copy number with respect to healthy controls.

Absolute quantification of extracellular alpha satellite DNA (ecASAT) repeats was performed by nanoplate-based digital PCR using few microliters of blood plasma from cancer patients as well as from BC negative controls. We found that copy number of ecASAT repeats is significantly higher in blood plasma of BC patients than in healthy controls.

The results suggest that the ec-hASAT copy number variation in human plasma could act as a reliable biomarker for early detection of BC.

## Introduction

Bladder cancer (in 90% of cases transitional cell tumour or urothelioma) is one of the tumours with the highest economic cost due to the extremely high recurrence rate and the resulting follow-up care. With almost world-wide 600,000 people affected and more than 200,000 deaths in 2020, it represents the tenth most diagnosed cancer in the world and statistics show these numbers are growing rapidly (Herranz et al. 2021, Sanli et al. 2017). Current methodologies for the diagnosis and follow-up care of BC are clearly suboptimal (Sanli et al. 2017; Zhu et al. 2019). Urinary cytology (to be carried out on at least 3 samples) has a sensitivity of 70% because it does not highlight tumours with a low degree of malignancy. Furthermore, it gives false positives in the case of bladder stones, and it cannot be implemented if there is an underlying inflammatory process, while giving uncertain results if previous therapeutic instillations have been performed. Cystoscopy, an invasive procedure, does not detect carcinoma in situ (CIS). Ultrasound can give false negatives because it does not explore the prostatic urethra, does not detect the Tis (intramucosal invasive) carcinoma and does not highlight carcinoma in situ (CIS) nor the smallest lesions. Furthermore, both magnetic resonance imaging (MRI) and computed tomography (CT) do not detect lesions smaller than a millimetre. When a tumour is diagnosed through these investigative methods, it is surgically removed via transurethral resection (TUR-BT). This endoscopic procedure is both therapeutic and diagnostic, as biopsy samples are analysed through histological examination. Unfortunately, histological results are not available during the preliminary evaluation and typically become accessible only after partial or total cystectomy. Consequently, the risk of incorrect therapeutic decisions is significant. This risk could be mitigated by the availability of a molecular test capable of rapidly predicting the presence or absence of the tumour during the preliminary clinical assessment. The results from the pathology division are diagnostic because through them it can be assign a “stage” and “grade” to the tumour. The “stage” of a cancer describes the size of the tumour and how far it has spread from where it first appeared. The “grade” (low or high) of a tumour depends on its microscopic appearance and how much the tumour cells differ in appearance from normal cells. It should be underlined that a tumour can be “high grade” even when small in size. For example, flat lesions CIS are always considered “high grade". This information is considered very important for selecting the most appropriate treatment. In any case based on the staging criteria, bladder cancer (BC) is presented in two main stages: non-muscle invasive (NMIBC) which is relatively confined to the bladder, with no signs of invasion of the underlying muscle as opposed to muscle invasive (MIBC) (Sanli et al. 2017). The NMIBC stage is more frequent and comprises about 70% of patients with bladder cancer (Herranz et al. 2021, Sanli et al. 2017).

The above-mentioned methods for detection of BC are not satisfactory for early diagnosis, and this makes it necessary to invest in the development of new, non-invasive and reliable biomarkers for this type of tumour which has high mortality rates. Currently, several liquid biopsy biomarkers are described that can be used to detect and follow the evolution of BC. These include sedimentary cells in urine, circulating tumour cells (CTC) in blood, RNAs and proteins in blood and urine, all of which, however, showed unsatisfactory sensitivity and specificity values (Lodewijk et al., 2018). In other words, BC biomarkers with excellent diagnostic performance and corresponding implementation data have not yet been identified and the current findings do not allow for firm recommendations of any of the identified biomarkers for use in the general population (Papavasiliou et al. 2023). Therefore, novel biomarkers showing promising results should be tested and further evaluated.

Circulating cell-free („ccf“, also called extracellular) nucleic acids (DNA, mRNAs, microRNAs or long non-coding RNAs), which refers to nucleic acid fragments found in the non-cellular components of liquid biopsies, were also identified as novel potential biomarkers (Herranz et al. 2021, Riethdorf et al. 2017, Lodewiyk et al. 2018, Tse et al. 2021, Huang et al. 2019). Higher levels of these circulating nucleic acid are detected in patients relative to healthy individuals (Corcoran et al. 2018, Tse et al. 2021, Huang et al. 2019). DNA from both nuclei and mitochondria seem to contribute to the whole extracellular DNA (ecDNA) pool but the biology of these DNA is still understudied regarding their sources, fragment length, stability and immunogenicity. However, the main sources of ecDNA are considered to be apoptosis and necrosis, even though both of them are not valid for satellite DNA (Bronkhorst, AJ et al 2016). An alternative source of ecDNA (especially satellite DNA) may be DNA secretion from a cell. Often, extracellular DNA (ecDNA) is associated with proteins, thus forming extracellular vesicles designated as exosomes. These proteins may protect the DNA from nuclease activity (Tutanov et al. 2022). Another approach was focused on the detection of mutations in circulating ecDNA (Sanli et al. 2017) or changes in epigenetic modifications such as DNA methylation (Hentschel et al. 2022; reviewed in Martinez et al. 2019). LINE-1 methylation status in ccfDNA combined with *Alu* ccfDNA integrity was used to discriminate patients with breast and lung cancer from healthy individuals (Park et al. 2014). Integrity of *Alu* repeat elements in ccfDNA was shown to be a prognostic marker for survival in primary and metastatic breast cancer (Madhawan et al. 2014; Cheng et al. 2018) while in prostate cancer, *Alu* ccfDNA integrity increases with disease severity and staging (Arko-Boham et al. 2019). Also, the length of telomeres in ccfDNA can serve as informative biomarker in many cancers (Wu et al. 2015; Dey et al. 2018; Shi et al. 2019). Considering satellite DNAs, they were shown to be the most overrepresented repeat in ccfDNA compared to their genomic abundance (Grabuscnig 2020). In addition, it was also shown that satellite DNA transcripts, which are overexpressed in different cancers, induce copy number increase of satellite repeats at pericentromeric regions via aberrant RNA:DNA hybrid formation (Bersani et al. 2015) and in this way could contribute to the enrichment of satellite repeats in ccfDNA of cancer patients. Increased levels of human satellite II DNA repeat circulating in the plasma of breast, gastric, lung and bile cancer patients as well as in those with sarcoma and Hodgkin’s lymphoma was detected and could be used as a potential diagnostic marker (Ozgur et al. 2021).

Unfortunately, despite the considerable effort invested in the aforementioned directions, no molecular marker has been introduced into clinical-diagnostic practice for the treatment of urological neoplasm.

In this paper, we characterized the involvement of highly repetitive DNA, in the form of circulating extracellular alpha satellite DNA (ccf-hASAT) in the plasma, as a powerful molecular marker for bladder cancer diagnostics. Satellite DNA is a highly repetitive DNA organized in a long array of tandem repeats and it is a fundamental component of the chromosomes of all eukaryotic species, forming megabase size arrays (Ugarković et al. 2022). For example, alpha satellite DNA (ASAT) is a repeat of 171 base pairs organized in tandem and forming the constitutive heterochromatin of all human chromosomes, mostly located at the centromeric and pericentromeric regions, representing at least 5–10% of the human genome (McNulty and Sullivan 2018).

Our results show that increased levels of circulating extracellular and chromosomal alpha satellite DNA in plasma are associated with the presence of bladder cancer when compared to healthy subjects. The alpha satellite thus represents a potential molecular marker of bladder cancer, and its quantification in the circulation could be of great help in diagnosis of specific tumour states as well as in testing the efficacy of the adopted therapeutic treatments. If these results are further confirmed on a greater number of cases, they will establish alpha satellite DNA as an absolute molecular marker for the diagnosis and prognosis of bladder cancer. Furthermore, it would pave the way for an in-depth, mechanistic study of the role that alpha satellite DNA plays in bladder cancer pathogenesis and disease progression.

## Materials and methods

### a) Sample collection and isolation of blood plasma circulating cell-free DNA

Blood plasma samples of bladder cancer patients as well as of healthy controls were provided by the Hospital of Polla, Salerno province, Italy. Informed consent was acquired from each participating individual before blood collection and the study has been authorized by the healthcare management on 08/03/2021. Half milliliter of blood of each bladder cancer patient and of healthy controls was collected in VACUETTE tubes for plasma extraction containing anticoagulant and processed immediately by centrifugation step at 5000 rpm for 5 minutes. The collected plasma was stored at -20 °C. Characteristics of patients and healthy controls with respect to age, gender and type of tumour are presented in Table 1.

The circulating ecDNA from plasma fraction was isolated by proteinase K digestion at 55 °C overnight and purified by QIAquick PCR Purification Kit (QIAGEN) according to manufacturer’s instructions.

### b) Absolute quantification of circulating cell-free DNA levels by digital PCR

Nanoplate based digital PCR (dPCR) procedure was performed using the QIAcuity 2-plex instrument (Qiagen, Hilden, Germany). The dPCR reaction mixture was assembled using QIAcuity 3X Eva Green PCR Master Mix, 10X primer mix (2 μM), RNase-free water and 1 μl of DNA template. After accurate vortexing, 12 μl of the above prepared mixture was transferred into the 24-well 8.5 K nanoplate and sealed with the nanoplate rubber seal. The sequences of primers used are: forward 5’-CACTCTTTTTGTAGAATCTGC-3’ reverse 5’- AATGCACACATCACAAAGAAG-3’. The 8.5 K nanoplate gives rise to 8500 single partitions in which the template is distributed randomly. The QIAcuity carries out fully automated processing including all necessary steps for plate priming, sealing of partitions, thermocycling, and image analysis. The amplification cycling protocol includes 95 °C for 2 min for enzyme activation and the following 40 cycles of 15 s at 95 °C for denaturation, 15 s at 60 °C for annealing, and 15 s at 72 °C for extension, and then a final step at 4°C. Fluorescence light is emitted only by positive partitions that contain a target molecule. The experiments were performed using also a negative control without target. Data were analyzed using the QIAcuity Suite Software V1.1.3 (Qiagen) and the results are expressed in copies of DNA/μL based on Poisson statistics analyses. The partitions produced by the machine resemble Poisson process since the targets end up in partitions independently and with a fixed rate. The Poisson distribution gives probabilities for positive integer random events. The parameter of this distribution, λ, is the expectation value for these events, which means it is the mean probability for a proportion of a counting process or the counting process for the dPCR analysis. Furthermore, the QIAcuity has embedded software that can quantify and produce reliable statistics. In our case, the statistical measure we considered was the Poisson confidence interval at a 95% level that, when plotted, shows whether the events differ with 95% confidence.

### d) Statistical analyses

The obtained dPCR results (Figure 1) were analyzed by Wilcoxon/Mann-Whitney U test which is used to analyze differences among two groups of samples: control and group of patients with bladder cancer.

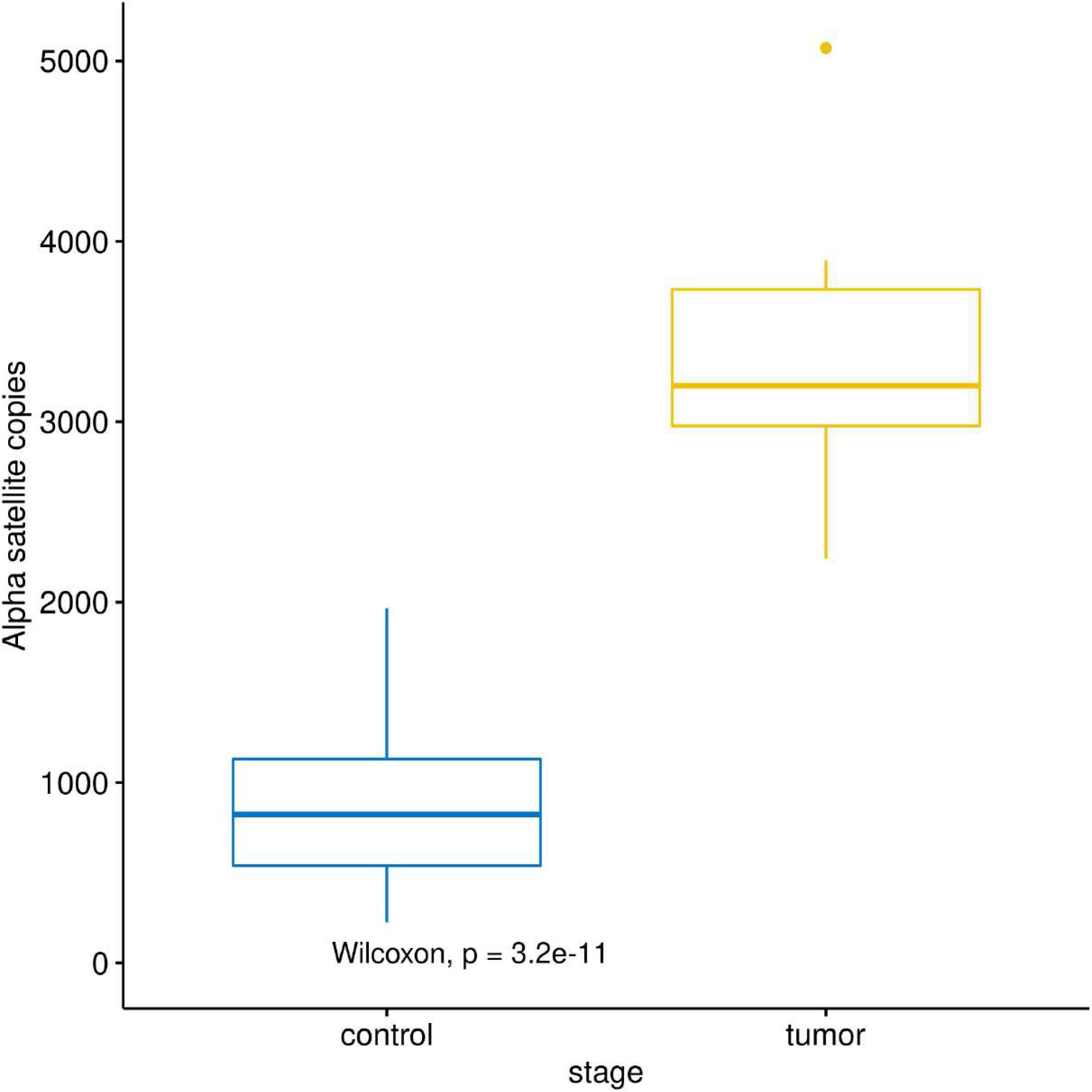

## Results

### 3.1. Structure and quantification of circulating cell-free alpha satellite DNA level in the blood plasma of bladder cancer patients

We isolated circulating cell-free DNA from blood plasma which was collected from 51 bladder cancer patients belonging to two main stages of the disease: non-muscle-invasive BC (NMIBC) and muscle-invasive BC (MIBC). Within each stage there were samples belonging to both low and high grade of disease. We also isolated ccfDNA from control groups which included 117 BC negative individuals and the number of samples of each group, gender, average age as well as age range is shown in Table 1. Proteinase K treatment was a crucial preliminary step in DNA purification from plasma to obtain positive results in PCR amplification. These findings suggest that extracellular DNA circulates in the form of a protein complex. Once purified, the amplicons from PCR amplification have shown to be of expected length size of 126 bp. Nanoplates-based digital PCR analysis was used to quantify the copy number of ccf alpha satellite DNA in the total ccf-DNA isolated from the blood plasma of patients belonging to two stages of BC as well as from healthy controls. All of the bladder cancer patients had copy number of ccf-alpha satellite DNA exceeding 2000 copies/μl plasma, whereas only 4 on 117 samples of negative controls exceeded this number (Table 1).

There was a significant difference in ccf alpha satellite DNA copy number between controls and BC groups of patients, however no significant difference was detected between MIBC and NMIBC samples. Also, no significant difference was observed between low and high-grade tumours belonging to either MIBC or NMIBC stage as well as between female and male samples. Based on these results, we suggest that the copy number of 2000 per μl of plasma could be taken as a threshold value between a negative and positive prediction of bladder cancer outcome. Hence, we may conclude that the extracellular level of alpha satellite DNA in human plasma can serve as a reliable biomarker for discriminating BC patients from healthy individuals. There was approximately only about 4 % of false positive, and more importantly, no false negative results, which is consistent with very favourable traits for a marker.

## Discussion

In this work, taking advantage of our long-time experience on satellite DNAs, we present the results demonstrating for the first time the possibility of using alpha satellite DNA as a plasma circulating cell-free molecular marker of bladder cancer. Various studies have highlighted that the altered expression of satellite DNAs can be associated with cancerous events (Ljubic et al 2022) and although overexpressed satellite DNA in different cancers could induce copy number increase of satellite repeats at pericentromeric regions via aberrant RNA:DNA hybrid formation (Bersani et al. 2015), no study has yet demonstrated that copy number variation of satellite DNA could be an excellent molecular marker capable of differentiating tumour states from the normal ones. These results have even greater relevance when we consider that the alteration can be diagnosed and quantified in liquid biopsies and therefore potentially enter clinical practice with the development of a rapid, reliable, sensitive, and non-invasive test for the diagnosis and/or prognosis of bladder cancer.

In our previous study we showed that the level of alpha satellite RNA is significantly increased in blood cells of metastatic prostate cancer patients and that it could serve as a marker for a specific stage of the disease (Ljubić et al. 2022). Here the results from plasma were obtained by testing 168 samples; 117 healthy controls and 51 cancer controls. All patients with bladder cancer showed to have higher copy numbers in circulation than healthy controls. Our results showed a significant increase of marker in cancer diagnosed patients compared to controls. The increased number of ccf alpha satellite DNA repeats was observed in both stages of bladder cancer (NMIBC and MIBC) as well as in low and high grade disease and could distinguish all groups of patients from healthy controls with high degree of certainty (p<10^−10^). ROC curves showed AUC value of 1.0 and demonstrated high specificity and sensitivity of ccf alpha satellite DNA repeat copy number as a potential diagnostic marker for bladder cancer. However, based on copy numbers of ccf alpha satellite DNA it was not possible to distinguish between NMIBC and MIBC, as well as between low and high grade of disease. Due to the copy number specificity of marker, which can distinguish with high precision between cancer patients compared to healthy controls, we proposed the potential use of this procedure as early diagnostic and predictive test for bladder cancer. The molecular marker proposed by us is not only able to precociously identify bladder cancer in its low-risk form but, being a quantitative marker, it is able to identify and quantify genomic instability at very early stage, when bladder cancer cannot be diagnosed with any other current method. Furthermore, it could be particularly effective in therapeutic follow-up care, giving a clear quantitative picture of the therapy efficiency.Some questions still remain unsolved. For example, why ASAT is detected in high copy number in circulation of cancer patients? Is ASAT circulating DNA bladder cancer specific? What is the copy number of the other satellite DNA in circulation as compared to hASAT? Is it possible to somehow treat the increased number of hASAT in cancer patients? To answer these interesting questions is not easy and it is not the aim of this paper but we will try to explain at least the origin of the high copy number variation of hASAT between samples through the mechanism of DNA repair acting on satellite DNA (Feliciello et al 2005, 2006). Satellite DNA is important to maintain the structure of the chromosome itself, therefore of the karyotype as well, while facilitating correct alignment of the homologous chromosomes as well as their proper separation during the various phases of mitosis and meiosis. Satellite DNA is under control of a specific DNA repair mechanism, which controls its evolution by means of a rolling-circle which amplifies single strand circular DNA (Feliciello et al 2006). The newly amplified satellite DNA repeats unit can be organized into heterochromatic state and the DNA repair mechanism is responsible also for the maintenance of the tandem organization in the genome. This mechanism was experimentally confirmed by showing for the first time the dynamic nature of satellite DNAs and their high copy number variations in a very small population, in a non-Mendelian inheritance (Feliciello et al 2005, 2006). These results have radically changed the paradigm about the molecular evolution of satellite DNA and demonstrated how important satellite DNA is in maintaining the karyotype and therefore the genome stability. Consistent with it, there are increasing evidences of satellite DNA alterations in the processes that lead to the development of cancer, such as variations in satellite DNA expression and cytogenetic abnormalities due to abnormal chromosomal recombination events. For example, different epithelial tumours, such as the pancreas, lung, kidney, colon and prostate cancers, abberant overexpression of satellite DNAs, including alpha satellite, was found (Ting et al. 2011; Ljubić et al. 2022) and increased levels of satellite transcripts can be detected as circulating cell-free satellite RNAs (Kishikawa et al. 2016). In addition, it is known that a significant portion of ccfDNA is composed of repetitive DNA sequences and some satellite DNA families, such as pericentromeric satellite II, are overrepresented in ccfDNA populations relative to their genomic abundance (Ozgur et al. 2021). However, the issue of determining satellite DNA copy number turnover in comparative analyses between normal and pathological states remains unsolved for long time. The limitation is of a technical nature, due to the lack of experimental procedures capable of precisely quantifying such a large number of copies of highly repetitive sequences.

The hypothesis upon which we based our research considered the turnover of the satellite DNA copy number and the presence of extracellular DNA (ecDNA) as consequences of genomic stability maintenance processes. Accordingly, genomic instability, as in cancer, should likely increase the production of extrachromosomal circular DNA in the cells as well as in circulation, especially in the plasma of cancer patients. The plasmatic ecDNA quantification should be quantified respect to the cellular form because it is expected to find ecDNA in a relatively low number of copies compared to those inside nuclei in the form of constitutive heterochromatin, and this assertion was confirmed by our study. However, more studies on different tumours are necessary in order to further validate the specificity of blood plasma ccf alpha satellite DNA as a molecular biomarker for bladder cancer diagnostics.

## Data Availability

All data produced in the present work are contained in the manuscript

## Acknowledgments

The authors would like to thank Carmela Iannibelli and Maria Giovanna Di Sevo from the “Luigi Curto” Hospital Facility in Polla, Italy, for their valuable contributions to this research. This work was supported by.

