## Supplementary material for "Extracellular alpha satellite DNA in human plasma as a novel molecular biomarker for early diagnosis of bladder cancer": Table 1

**Table 1.** Two Groups of individuals (healthy and bladder cancer patients) used in this study with the following characteristics: number of samples (N); number of females/males; age average (years) with minimal and maximal age; average and minimal and maximal number of copy/µl of hASAT biomarker.

| | **Number of samples** | **Female/Male** | **Age average (years) min/max** | **hASAT average (copy/µl) min/max** |
|---|---|---|---|---|
| **Healthy controls** | **117** | **69/48** | **58**<br>**21/88** | **683**<br>**108/1965** |
| **Bladder cancer patients** | **51** | **8/43** | **72**<br>**45/90** | **6584**<br>**1979/19007** |
